# Peer advocacy and access to hospital care for people who are homeless in London, United Kingdom 2019-2023: a cohort study

**DOI:** 10.1101/2025.09.11.25335486

**Authors:** Lucy Platt, Andrew Guise, Paniz Hosseini, Kate Bowgett, Mani Cudjoe, PJ Annand, Michael “Spike” Hudson, Maame Esi D. Yankah, Dee Menezes, Rosa Legood, Rob Aldridge, Andrew Hayward, Serena Luchenski, Elizabeth Williamson, Sujit D Rathod

**Affiliations:** Faculty of Public Health and Policy, London School of Hygiene and Tropical Medicine, 15-17 Tavistock Place, London WC1H9SH, United Kingdom, United Kingdom; Population Health Sciences, King’s College London, Addison House, London SE11UL, United Kingdom; Groundswell, 3 Brixton Rd, London SW9 6DE, United Kingdom; Centre for Care, University of Sheffield, Social Research Institutes, The Wave, Sheffield, S102AH, United Kingdom; Institute of Health Informatics Research, University College London, 222 Euston Road, London, NW1 2DA, United Kingdom; Institute of Epidemiology and Healthcare, University College London,□Gower Street,□London,□WC1E 6BT, United Kingdom; Faculty of Epidemiology and Population Health, London School of Hygiene and Tropical Medicine, Keppel Street, London WC1E7HT, United Kingdom

**Keywords:** Homelessness, peer support, hospital engagement, evaluation, cohort study

## Abstract

**Objectives:** To measures differences in hospital use between homeless adults using a peer advocacy service (clients) and non-clients in London.

**Design:** We conducted a cohort study with linkage to hospital episode statistics (HES) one year prior and post enrolment.

**Setting:** London, UK

**Population:** People who are homeless in London aged over 18 years residing in a hostel, attending a day centre, or being referred by a homelessness service; experiencing difficulties accessing healthcare; and speaking either English or Polish. Participants were required to provide consent for linkage to Hospital Episode Statistics (HES). To be classified as a client, individuals must have used the HHPA service at least once between January and July 2021; non-clients were those who had never used the service.

**Intervention:** Peer advocacy is the provision of support by trained advocates with lived experience of homelessness to individuals to overcome barriers to accessing health services.

**Outcomes:** The primary outcome was not attending a scheduled outpatient appointment (‘did-not-attend’, DNA) over 12 months post-recruitment, commencing from their baseline interview date. Secondary outcomes included the number of A&E and inpatient admissions (all and planned admissions) during that same period.

**Methods:** We estimated the probability of DNA using Poisson regression and the number of inpatient admissions and accident and emergency visits using linear regression models. Models adjusted for: i) propensity score weights; and ii) propensity scores and imbalanced confounders. Sensitivity analyses assumed that participants who did not link to HES had no hospital attendance. Secondary analyses examined differential effects by type of peer advocacy engagement (new vs. ongoing clients; supported vs. unsupported) and anxiety or depression symptom scores measured with the Public Health Questionnaire 4 (PHQ4).

**Results:** 153 clients and 159 non-clients were recruited between July-December 2021. Most were male (77.5%) with median age of 48 years. Weighted regression models suggested no effect of peer advocacy on DNA(Rate Ratio (RR) 0.97 95% CI 0.67, 1.42), no difference in the mean number of A&E visits (0.86, 95% CI -0.06, 1.79) but a difference in inpatient admissions 1.14 (95% CI 0.52,1.75). Sensitivity analyses suggested a higher number of completed outpatient appointments (1.77 95% CI 0.13,3.40) among clients. Clients with PHQ4 scores of 9-12 had greater probability of DNA at outpatient appointments (RR 1.98 95% CI 1.0,3.89), those with scores of 6-8 had 3.38 (95% CI 0.05,6.71) more completed appointments and 1.09 (95% CI 0.56-1.63) more inpatient admissions, relative to non-clients.

**Conclusions:** In the context of COVID-related disruptions to the work of peer advocates and health services we found mixed evidence on the effect of peer advocacy: with no impact on outpatient appointments or use of emergency services; but increased inpatient admissions.

**Article Summary:** *Strengths and Limitations of this study:*

- Use of linked hospital data minimised recall bias by relying on objective outcome measures sourced from national Hospital Episode Statistics (HES), enhancing data accuracy and reliability.
- Application of propensity score weighting addressed baseline imbalances between intervention and comparison groups, allowing more robust adjustment for confounding and strengthening causal inference in a non-randomised design.
- Potential for residual confounding remains, given notable differences between peer advocacy clients and non-clients and the possibility of unmeasured variables (e.g. severity of illness or motivation to seek care) influencing outcomes.
- Data linkage was incomplete, with 26% of participants not matched to HES records, introducing potential selection bias and limiting confidence in sensitivity analyses that assumed non-attendance among unlinked individuals.
- Impact of COVID-19 disrupted both intervention fidelity and usual care, with reduced in-person peer support and altered healthcare access potentially diluting the observed effects of the intervention and affecting generalisability.

## Introduction

Homelessness includes sleeping outside, in hostels, shelters and temporary accommodation, as well as residing in unsafe and inadequate housing (e.g. overcrowded conditions or settings involving domestic violence).(1) In the UK, all forms of homelessness are increasing. In 2023, 3898 people were observed sleeping outside on a single night - a 27% increase from 2022 - and this is thought to underestimate the true number.(2) Local authorities estimated a 20% increase in households in temporary accommodation between 2018-19 and 2022.(3)

Homeless organisations in England and Wales report that 14% of homeless people have chronic respiratory disease, 17% have tuberculosis, and 10% are living with hepatitis C infection. Overall, 80% report one or more chronic physical health problems.(4) Self-reported mental health diagnoses increased from 45% in 2014 to 82% in 2022.(4, 5) Standardised mortality rates among marginalised populations - including those with experience of homelessness-are between 7.9 and 11.9 times higher for men and women compared with stably-housed populations, and four times higher than populations in the most deprived areas.(6)

These inequalities in health reflect not only multiple experiences of exclusion and elevated health needs, but also limited access to healthcare. People experiencing homelessness have high rates of accident and emergency (A&E) use (50% at least once in the past year) and hospital admissions (38% in the last year). This represents avoidable ill-health and distress, as week as considerable costs to the health system: healthcare costs for homeless individuals are estimated to be eight times higher than those of the general population.(7)

Peer support can address several healthcare challenges associated with homelessness. The availability of a trusted advocate - with lived experience of homelessness - can reduce structural (e.g. stigma or alienation, transport), social (e.g navigating interactions with staff) and bureaucratic (e.g. paperwork) barriers to accessing healthcare.(8-12) Evidence supports the effectiveness of peer support in mental health settings, (13) in the context of drug use (14-16) and highlights its potential among sex worker, prison and homeless populations.(12, 17, 18) However, peer support in the UK has historically received limited funding and lacks national government backing.(1) Moreover, a shortage of evaluations using well-theorised interventions and rigorous study designs means there is still insufficient evidence to support a more comprehensive roll-out.(13, 19)

We conducted a mixed-methods evaluation of a peer advocacy intervention among homeless adults in London, assessing its impact, cost-consequences and processes related to healthcare utilisation.(20) This paper reports findings from the impact evaluation component..

## Methods

### Study design

We conducted a prospective cohort study of people experiencing homelessness who had difficulty meeting their healthcare needs: both peer advocacy clients and non-clients.

### Patient and Public Involvement

We used participatory approaches, involving people with lived experience of homelessness or those working with homeless individuals as co-researchers, to develop methods, gather data, and conduct analysis. The project was overseen by a steering group consisting of people working in the homeless sector and with lived experience of homelessness to guide the design and interpretation of findings.(20)

### Intervention and impact of COVID-19

Peer advocacy was defined as the provision of support by trained advocates - ‘peers’ with lived experience of homelessness - to individuals currently experiencing homelessness, with the aim of overcoming barriers to accessing health services. We evaluated the Homeless Health Peer Advocacy (HHPA) service, delivered by Groundswell, a non-governmental organisation working alongside specialist homelessness services in London.

HHPA supports people who are homeless and have difficulty managing their healthcare needs. For the purposes of this study, homelessness was defined as residence in a hostel, attendance at a day centre, or referral by a homelessness service. Support includes assistance with healthcare appointments. Clients are generally aged over 25, although HHPA also works with individuals aged 18 to 24 years. Peers receive training, regular supervision, and support for their own personal development, and are reimbursed for expenses. Initial meetings allow peers and clients to build rapport. Support is not time-bound and may be open-ended where needed (21)

Support activities include accompaniment to healthcare appointments, assistance with GP registration, one-on-one conversations, and financial help with transport. Peers also signpost clients to other relevant services when needs fall outside HHPA’s remit (e.g. nutrition, housing, or legal advice). Clients may self-refer, be referred by a key worker, or be approached through peer ‘in-reach’ activities at hostels and day centres. HHPA operated in 10 of London’s 32 boroughs serving approximately 600 clients annually.

During the recruitment period (July to December 2021), HHPA was providing less direct in-person advocacy, instead conducting welfare calls by phone, joining clients in online consultation for outpatient appointments and offering increased support for independent travel. Although 21 peer advocates were available, there were working less frequently. At the same time, the NHS was experiencing significant delays in outpatient care and planned inpatient admissions. Simultaneously, London’s outdoor sleeping population was provided emergency accommodation in hotels, where GP registration and other health and social care services were facilitated.(22) Despite these disruptions, the research team deemed HHPA to be functioning adequately to warrant continuation of the evaluation..

### Eligibility criteria

Individuals aged over 18 years were eligible if they met the following criteria: residing in a hostel, attending a day centre, or being referred by a homelessness service; experiencing difficulties accessing healthcare; and speaking either English or Polish. Participants were required to provide consent for linkage to Hospital Episode Statistics (HES). To be classified as a client, individuals must have used the HHPA service at least once between January and July 2021; non-clients were those who had never used the service.

### Recruitment

Clients were recruited by peer advocates. Non-clients were recruited concurrently and opportunistically from hostels and day centres in areas of London where Groundswell was not commissioned to operate the HHPA service and where individuals were receiving the usual available support services. All participants received a copy of the Pavement which lists available services, and were reimbursed £10.

### Baseline Data collection

Peer advocates and co-researchers invited clients and non-clients, respectively, to complete a structured questionnaire (in English or Polish) on a tablet (Open Data Kit V.1.28.4) or administered in person, with 17 conducted by phone. Indicators were drawn from validated measures and other surveys (4, 23-25) and embedded in a linked qualitative study.(26-28) Questions included demographic characteristics; indicators of multiple exclusion health; mental and physical health issues; health and social service use and barriers; and alcohol and other drug use. We measured anxiety and depression using the Patient Health Questionnaire (PHQ)-4 (29) categorising participants with scores of 0-5 as low risk, 6-8 as moderate risk and 9-12 as severe risk.(30) (see supplementary file for the full questionnaire). Personal identifiers were used to link participants to three HES databases (England-wide) to extract attendance at scheduled outpatient appointments, use of A&E and inpatient admissions for one year prior to and post baseline interview. Analyses focussed on linked participants whose records were identified in any one of these six databases.

### Outcomes

The primary outcome was not attending a scheduled outpatient appointment (‘did-not-attend’, DNA) over 12 months post-recruitment, commencing from their baseline interview date. Secondary outcomes included the number of A&E and inpatient admissions (all and planned admissions) during that same period.

### Confounders

Pre-specified confounders included: age; gender (male, female, other (including non-binary, genderqueer, agender or gender fluid)); ethnicity (Black, Asian, multiple, White); time since first episode of homelessness and; citizenship (UK vs non-UK). Potential confounders included: historic indicators of exclusion (past injecting drug use, imprisonment, and begging (asked passers-by for money in a public place)); current indicators of exclusion (location for sleeping last night categorised as stable (hostel, supported housing, own tenancy) and non-stable (rough, sofa surfing, friends, emergency, bed and breakfast accommodation); use of heroin in the last 12 months); reporting transportation as a barrier to healthcare; and symptoms of anxiety or depression.

### Missing data

Primary analyses assumed participants with at least one record in any of the six HES datasets in the 12 months pre or post-recruitment had zero visits for their other datasets for which records were not found. Those who were not identified in any of the databases were assumed to be unlinked and missing. Sensitivity analyses assumed that anyone not linked to any of the six HES databases did not have a scheduled outpatient appointment, or attend it, nor attended A&E, nor had an inpatient admission during the12 month follow-up and were included in the analysis.

### Statistical analyses

We compared characteristics of clients and non-clients and linked and unlinked participants using medians and interquartile ranges for continuous data and percentages for categorical data, and with standardised differences (for linked-unlinked comparisons).

We estimated the relative probability of DNA at an outpatient appointment using a weighted Poisson regression model, among participants who had ≥1 appointments scheduled in the follow-up period. Models are offset for the number of appointments in that time. We used linear regression models to estimate the difference in number of DNAs, attended appointments, A&E visits and inpatient admissions.

Firstly we present univariable associations for all outcomes. Secondly, potential confounders were included where there was a standardised difference >0.1 for linked clients compared to linked non-clients at baseline.(20) To account for confounding, we calculated propensity scores in a logistic regression model where the outcome was whether the participant was a peer advocacy client. We combined similar categories with small strata (n<10) and considered interaction terms to better fit this model. With the final propensity score model we calculated the stabilised inverse probability of treatment weights, and re-calculated the standardised differences with these weights. We defined variables as persistently imbalanced where the weighted standardised difference was >0.1 (Table S1) We weighted the regression analyses with the inverse probabilities.(20) Thirdly, given the possibility of residual confounding after propensity score weighting, we repeated the weighted regression analyses including the persistently imbalanced variables as covariates.

### Sensitivity and secondary analyses

As a sensitivity analysis we imputed a ‘0’ outcome for A&E, inpatient and completed outpatient counts for participants who did not link to any HES dataset. Secondary analyses focussed on repeating the weighted models by their baseline depression-anxiety symptom score, through the PHQ-4 tool. We also examine differences between type of engagement with HHPA. This included examining differences between ‘new’ clients defined as those who had 0-1 engagements (any type) versus ‘on-going’ clients defined as those ≥2 engagements scheduled with the peer advocacy programme at the point of study recruitment. To examine the reduced service that HHPA was conducting we also examined differences between clients who had ≥2 peer-supported contacts consisting of a welfare visit, support with GP registration and accompaniment to a health care (hospital, GP, dentist) appointment (termed supported clients) compared to those who had either 0 or 1 contacts (termed unsupported) and mostly using the programme for assistance with travel during the study observation period.

### Ethics

Informed consent on participation in the cohort study, the collection of personal identifiers and linking to hospital episode statistics was obtained from all participants. The evaluation was approved by the London School of Hygiene and Tropical Medicine (Ref: 18021) and the Dulwich Research Ethics Committee (IRAS 271312).

## Results

A total of 311 participants were recruited, 153 HHPA clients (35.2%, n=434) and 158 non-clients (91.3% n=164 approached). Overall, 77.5% were male and the median age was 48 years. Compared to non-client, clients were older (50 years, IQR 43-58 vs. 45 years, IQR 39-55) and had a longer duration of homelessness (20 years, IQR 7-33 vs 9 years IQR 3-17). Fewer clients had slept outside the previous night compared to non-clients (1.3% vs13.9%) but they were more likely to have past experience of injecting drugs (40.5% vs. 17.7%), begging (51.0% vs 31.6%) or incarceration (58.8% vs 41.1%) and used heroin in the last year (29.2% vs 19.0%) compared to non-clients. Across both groups, 38% had severe symptoms of depression or anxiety (Table 1).

**Table 1.** Characteristics of all participants and by use of peer advocacy intervention.

|  | HHPA clients (n=153) | Non-clients (n=158) | Total (n=311) |
| --- | --- | --- | --- |
|  | n (col %) | n (col %) | n (col %) |
| Gender |  |  |  |
| Male | 118 (77.1) | 126 (79.7) | 244 (78.5) |
| Female | 35 (22.8) | 32 (20.2) | 67 (21.5) |
| Non-binary, genderqueer/fluid, agender | <5 | <5 | <5 |
| Age, years (median, IQR) | 50 (43, 58) | 45 (39, 55) | 48 (40, 57) |
| Sexual orientation |  |  |  |
| Heterosexual | 138 (92.0) | 132 (83.5) | 270 (86.8) |
| Gay, lesbian, bisexual, Other | 15 (9.9) | 25 (15.8) | 41 (13.2) |
| Ethnicity |  |  |  |
| White only | 106 (69.3) | 97 (61.4) | 203 (65.3) |
| Black/Black British only | 12 (7.8) | 15 (9.5) | 27 (8.7) |
| Asian, Other, multiple, refuse | 35 (22.9) | 46 (29.1) | 81 (26.0) |
| Citizenship |  |  |  |
| Other | 26 (17.0) | 42 (26.5) | 68 (21.9) |
| British | 127 (83.0) | 116 (73.4) | 243 (78.1) |
| Education completed |  |  |  |
| Less than secondary | 18 (11.8) | 22 (14.2) | 40 (13.0) |
| Secondary | 88 (57.9) | 73 (47.1) | 161 (52.4) |
| More than secondary | 46 (30.3) | 60 (38.7) | 106 (34.5) |
| Duration homeless years (median, IQR) | 20 (7, 33) | 9 (3, 17) | 11 (5, 28) |
| Sleeping location, last night |  |  |  |
| Slept rough / public location | 2 (1.3) | 22 (13.9) | 24 (7.7) |
| Hostel | 104 (68.0) | 114 (72.1) | 218 (70.1) |
| Own tenancy | 27 (17.6) | 5 (3.2) | 32 (10.3) |
| Other | 20 (13.1) | 17 (10.8) | 37 (11.9) |
| Past experience of multiple exclusion |  |  |  |
| Begged | 78 (51.0) | 50 (31.6) | 128 (41.2) |
| Sold sex | 15 (9.8) | 15 (9.5) | 30 (9.6) |
| Incarcerated | 90 (58.8) | 65 (41.1) | 155 (49.8) |
| Injected drugs | 62 (40.5) | 28 (17.7) | 90 (28.4) |
| Healthcare barriers, current <sup>a</sup> |  |  |  |
| Problems with transportation | 105 (68.6) | 83 (52.5) | 188 (60.5) |
| Uncertainty about place and provider | 96 (62.7) | 89 (56.3) | 185 (59.5) |
| Couldn't get appointment | 87 (56.9) | 74 (46.8) | 161 (51.8) |
| Used heroin in last 12 months |  |  |  |
| No | 92 (60.1) | 128 (81.0) | 220 (70.7) |

|  |  | **HHPA clients (n=153)** | **Non-clients (n=158)** | **Total (n=311)** |
| --- | --- | --- | --- | --- |
|  | Yes | 61 (40.0) | 30 (19.0) | 91 (29.2) |
| PHQ4 score category |  |  |  |  |
|  | 0-5 (Low) | 55 (36.9) | 57 (36.8) | 112 (36.8) |
|  | 6-8 (Moderate) | 37 (24.8) | 39 (25.2) | 76 (25.0) |
|  | 9-12 (Severe) | 57 (38.3) | 59 (38.1) | 116 (38.2) |

### Characteristics of participants linked into Hospital Episode Statistics

Overall 229 participants (74%) were linked to least one record in a HES dataset (outpatient, inpatient, or A&E) in the 12 months either pre or post-enrolment. A total of 82 participants (26.4%) had no records located in any dataset and were unlinked. Among clients, 84% were linked to a HES record compared to 63% of non-clients. A total of 174 participants had ≥1 attendances at an outpatient appointment, 196 had used A&E services and 126 had an inpatient admission. Table 2 summarises the recruitment and data linkage of peer advocacy clients and non-clients. Subgroups less likely to link included those identifying as Asian/multiple/other ethnicity (34.2%), those with non-UK citizenship (30.5%), those with less than secondary school education (18.3%), and those reporting an unstable sleeping location last night (26.8%). The differences between linked peer advocacy clients and linked non-clients mirrored differences within the total sample (Supplementary Table 1)

**Table 2.**
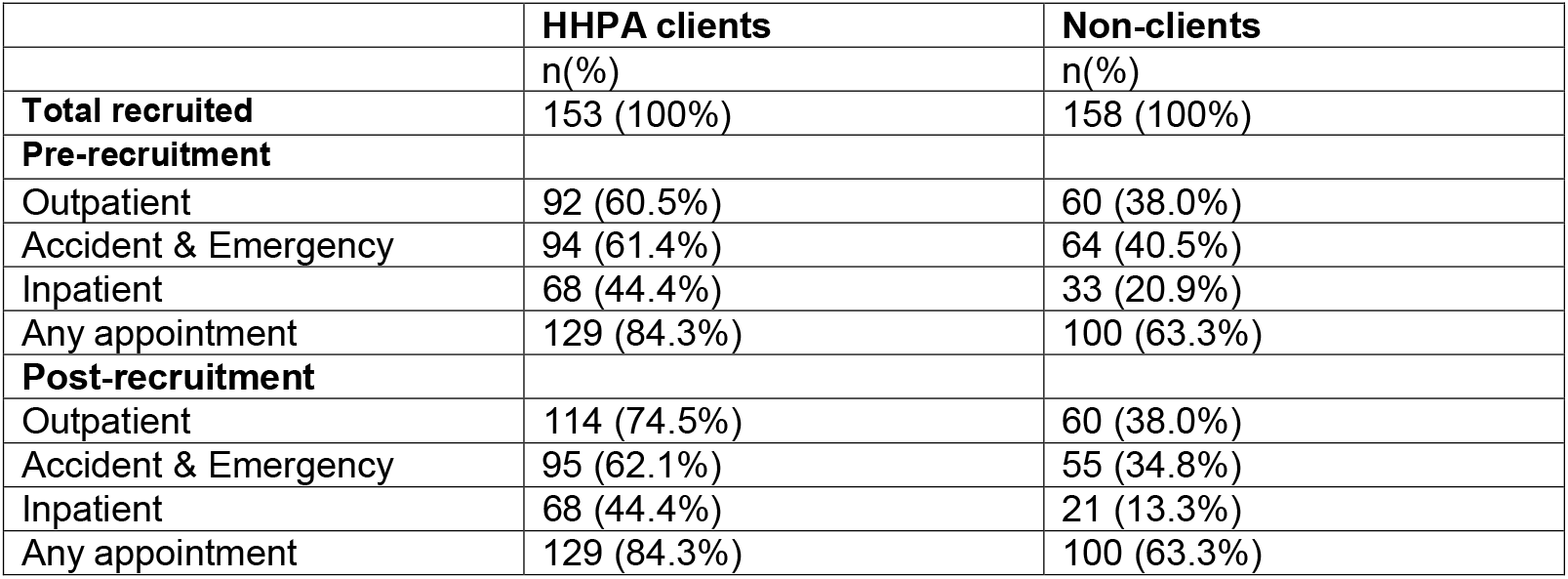
Linkage of cohort study participants with Hospital Episode Statistics records for the 12 months pre-and post-recruitment, London, UK.

### Hospital attendance among linked participants

For both clients and non-clients the median number of scheduled and completed outpatient appointments was higher among participants at 12 months pre-recruitment compared to 12 months post-recruitment (Table 3). The number of A&E visits was slightly higher among HHPA clients pre-recruitment compared to post-recruitment but the same for non-clients. Between arms there were large differences across all the hospital outcome measures both before and after study recruitment. Non-clients had one scheduled outpatient appointments in the 12 months before recruitment, compared to four appointments for clients (SD=0.50) and of these appointments, 0.0 ended in a DNA for non-clients compared to one for clients (SD=0.45). After generating stabilised inverse probability of treatment weights, the only confounding variables where the weighted standardised differences remained >0.10 were the hospital use variables for the 12 months before study recruitment and years since first episode of homelessness. Variables hypothesised as potential confounders which had weighted standardised differences <0.10 were not included in the analysis or reported here.

**Table 3.**
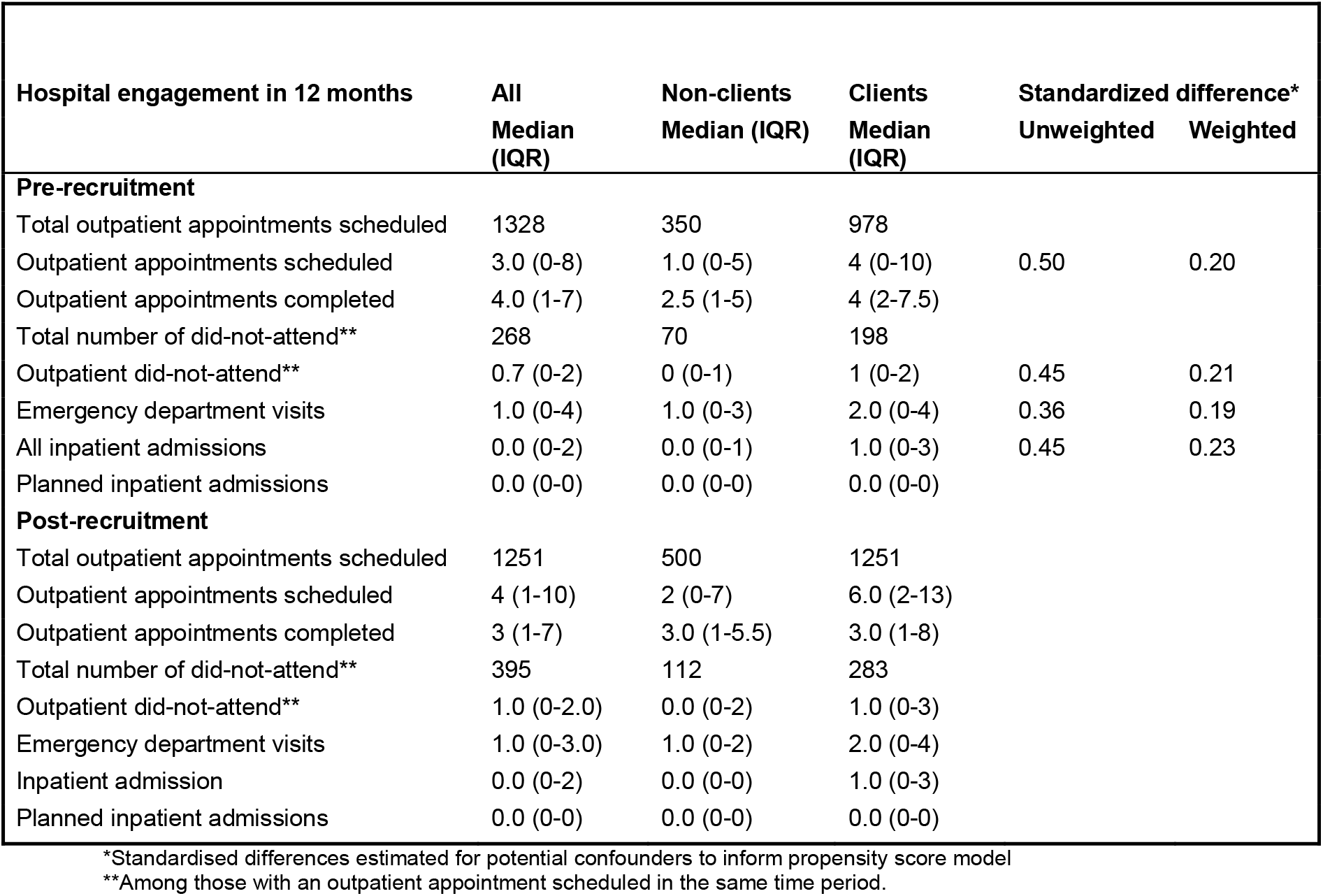
Hospital engagements in the last 12 months pre and post-recruitment for linked participants (n=229) and by intervention arm.

### Effect of peer advocacy

In weighted regression model (Table 4), we found no evidence of a difference between clients and non-clients in the probability of a DNA at an outpatient appointment in the 12 months post-recruitment (rate ratio (RR) 0.97, 95% CI 0.67, 1.42). The RR was similar in the weighted model adjusted for imbalanced confounders (RR 1.03 95% CI 0.69,1.54). We found no evidence of difference in the number of DNA at outpatient appointments, the number of completed outpatient appointments, the number of A&E visits, or planned inpatient admissions between arms. In a weighted linear regression model we did find evidence that, relative to non-clients, clients had 1.14 more total inpatient admissions (including both planned and unplanned) (95% CI 0.52, 1.75) in the 12 months after study recruitment, with a similar finding from the weighted multivariable model.

**Table 4.** Effect of being a peer advocacy client on hospital use among HES-linked cohort study participants who are homeless in London, United Kingdom, 2021-2022.

| Outcome (Peers vs non-clients) | Unadjusted | Weighted<sup>a</sup> | Weighted and adjusted<sup>b</sup> |
| --- | --- | --- | --- |
| **Model including linked Participants only** |  |  |  |
|  | Estimated rate ratio (95% confidence interval) |  |  |
| Did-not-attend for outpatient appointment | 1.01 (0.75, 1.35) | 0.97 (0.67, 1.42) | 1.03 (0.69, 1.54) |
|  | Estimated mean difference (95% confidence interval) |  |  |
| Number of DNA outpatient appointments | 0.62 (-0.18, 1.42) | 0.45 (-0.34, 1.25) | 0.38 (-0.46, 1.23) |
| Number of completed outpatient appointments | 1.26 (-1.09, 3.61) | 0.95 (-1.74, 3.63) | 0.78 (-2.00, 3.55) |
| Number of A&E visits | 1.22 (0.16, 2.29) | 0.86 (-0.06, 1.79) | 0.53 (-0.30, 1.36) |
| Number of inpatient admissions | 1.51 (0.73, 2.30) | 1.14 (0.52, 1.75) | 0.94 (0.38, 1.52) |
| Number of planned inpatient admissions | 0.44 (-0.10, 0.98) | 0.30 (-0.04, 0.65) | 0.24 (-0.03, 0.52) |
| **Sensitivity analysis with all participants** |  |  |  |
|  | Estimated mean difference (95% confidence interval) |  |  |
| Number of DNA outpatient appointments | 1.14 (0.66, 1.62) | 0.68 (0.13, 1.23) | 0.50 (-0.06, 1.02) |
| Number of completed outpatient appointments | 2.80 (1.43, 4.18) | 1.77 (0.13, 3.40) | 1.25 (-0.40, 2.91) |
| Number of A&E visits | 1.44 (0.63, 2.25) | 0.82 (0.14, 1.50) | 0.47 (-0.15, 1.08) |
| Number of inpatient admissions | 1.38 (0.80, 1.97) | 0.94 (0.41, 1.47) | 0.76 (0.27, 1.25) |
| Number of planned inpatient admissions | 0.40 (0.00, 0.80) | 0.25 (0.00, 0.50) | 0.19 (-0.01, 0.39) |

<sup>a</sup>Weights account for education, gender, sexual identity, ethnicity, age, duration of homelessness, history of begging, injecting drug use, incarceration, location sleeping last night, transportation problems, PHQ4 symptoms, use of heroin in the past 12 months and number of outpatient appointment, number of DNA and number of A&E visits scheduled in 12 months before study recruitment
<sup>b</sup> Adjusted for years since first homeless and hospital use variables for the 12 months before study recruitment (i.e. number of scheduled outpatient visits, number of did-not-attend outpatient visits, number of emergency department visits, number of inpatient admissions).

### Sensitivity and secondary analyses

In sensitivity analyses the number of completed outpatient appointments was higher among clients in the weighted model (1.77 95% CI 0.13, 3.40). The number of DNA outpatient appointments was also higher (0.68 85% CI 0.13,1.23) among clients compared to non-clients. The number of inpatient admissions (all) was higher among clients, and we observed 0.25 (95% CI 0.0-0.50) more planned inpatient admissions and 0.82 (95% CI 0.14,1.50) more A&E visits among clients compared to non-clients (Table 4).

There were 55 clients and 57 non-clients graded low risk for anxiety or depression; 37 clients and 39 non-clients were categorised as moderate and 57 clients and 59 non-clients as severe (Table 5). Clients categorised as severe had 1.98 (95% CI 1.00-3.89) higher risk of DNA at an outpatient appointment compared to non-clients. Clients with moderate symptoms had 3.38 (95% CI 0.05-6.71) more completed outpatient appointments compared to non-clients. They also had 1.09 (95% CI 0.56-1.63) more inpatient admissions and 0.52 (95% CI 0.12-0.92) planned inpatient admissions than non-clients.

**Table 5.**
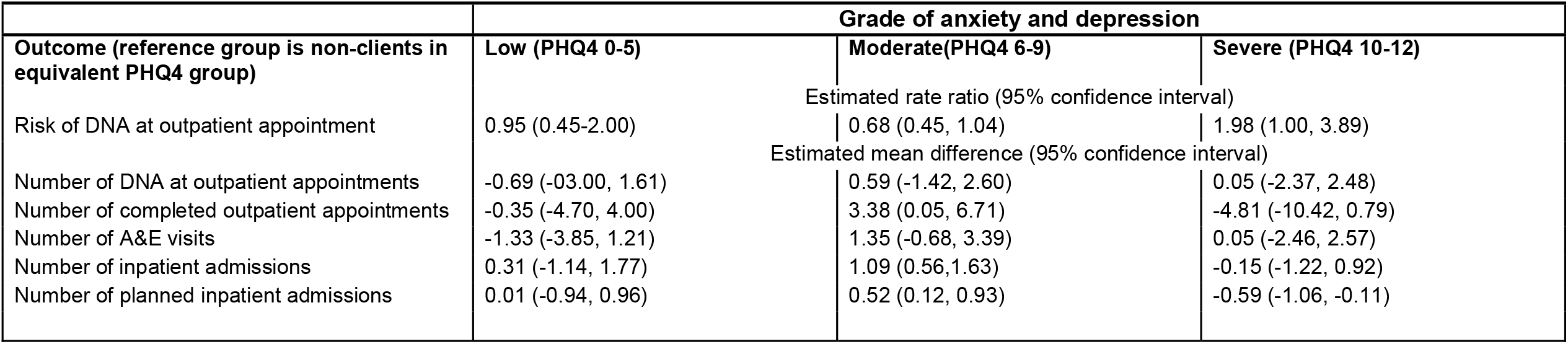
Secondary Analyses: Effect of peer advocacy on hospital use within categories of anxiety and depression symptoms, London, United Kingdom (2021-2022)

| Outcome (reference group is non-clients in equivalent PHQ4 group) | Grade of anxiety and depression |  |  |
| --- | --- | --- | --- |
|  | Low (PHQ4 0-5) | Moderate(PHQ4 6-9) | Severe (PHQ4 10-12) |
|  | Estimated rate ratio (95% confidence interval) |  |  |
| Risk of DNA at outpatient appointment | 0.95 (0.45-2.00) | 0.68 (0.45, 1.04) | 1.98 (1.00, 3.89) |
|  | Estimated mean difference (95% confidence interval) |  |  |
| Number of DNA at outpatient appointments | -0.69 (-03.00, 1.61) | 0.59 (-1.42, 2.60) | 0.05 (-2.37, 2.48) |
| Number of completed outpatient appointments | -0.35 (-4.70, 4.00) | 3.38 (0.05, 6.71) | -4.81 (-10.42, 0.79) |
| Number of A&E visits | -1.33 (-3.85, 1.21) | 1.35 (-0.68, 3.39) | 0.05 (-2.46, 2.57) |
| Number of inpatient admissions | 0.31 (-1.14, 1.77) | 1.09 (0.56, 1.63) | -0.15 (-1.22, 0.92) |
| Number of planned inpatient admissions | 0.01 (-0.94, 0.96) | 0.52 (0.12, 0.93) | -0.59 (-1.06, -0.11) |

Overall, 80 (52.3%) were new clients and 73 (47.7%) were ongoing clients. There were 86 (56.2%) unsupported and 67 (43.8%) supported clients. We repeated the weighted regression analyses, for comparison of these groups against non-clients (Table 6). There were few differences across any of the outcomes with the exception of use of A&E and in-patient admissions. Relative to non-clients, new peer advocacy clients had 0.75 more inpatient admissions (95% CI 0.14-1.36) and on-going clients had 1.69 more admissions (95% CI 0.55-2.83). Relative to non-clients, non-supported clients had 1.06 (95% CI 0.32-1.79) more in-patient admissions and supported peer advocacy clients had 1.31 (95% CI 0.30-2.31). Finally supported clients had 1.19 (95% CI 0.03-2.36) more A&E visits relative to non-clients.

**Table 6.** Secondary Analyses: Effect of being a peer advocacy client on hospital use within client subgroups, London, United Kingdom, 2021-2022.

| Outcome (reference group= non clients) | New and on-going clients |  | Supported vs unsupported clients |  |
| --- | --- | --- | --- | --- |
|  | New clients (n=80) | On-going clients (n=73) | Non-supported clients (n=86) | Supported clients (n=67) |
|  | Estimated rate ratio (95% confidence interval) |  |  |  |
| Risk of DNA at outpatient appointment | 1.03 (0.69-1.5) | 0.90 (0.58-1.39) | 1.02 (0.68, 1.52) | 0.90 (0.58, 1.39) |
|  | Estimated mean difference (95% confidence interval) |  |  |  |
| Number of DNA at outpatient appointments | 0.71 (-0.34, 1.77) | 0.00 (-0.91, 0.92) | 0.58 (-0.48, 1.64) | 0.08 (-0.78, 0.94) |
| Number of completed outpatient appointments | 1.40 (-1.54, 4.34) | 0.45 (-2.32, 3.22) | 1.04 (-1.86, 3.94) | 0.84 (-1.92, 3.61) |
| Number of A&E visits | 0.44 (-0.55, 1.43) | 1.47 (0.07, 2.86) | 0.69 (-0.41, 1.79) | 1.19 (0.03, 2.36) |
| Number of inpatient admissions | 0.75 (0.14, 1.36) | 1.69 (0.55, 2.83) | 1.06 (0.32, 1.79) | 1.31 (0.30, 2.31) |
| Number of planned inpatient admissions | 0.15 (-0.09, 0.39) | 0.54 (-0.21, 1.29) | 0.13 (-0.09, 0.37) | 0.61 (-0.22, 1.44) |

## Discussion

This study aimed to measure the effectiveness of peer advocacy on hospital use among people who are homeless in London, conducted in the context of disruptions to both HHPA and healthcare appointments due to COVID-19. Results were mixed. While we did not find evidence of an effect of peer advocacy on the number of DNAs at outpatient appointments or use of A&E, peer advocacy clients had 1.14 more total inpatient admissions in the 12 months after study recruitment relative to non-clients. Sensitivity analysis suggested clients had 0.94 more inpatient admissions; 0.82 more A&E visits,1.77 more completed outpatient appointments, but 0.68 more DNAs. We found that the effectiveness of peer advocacy is linked to symptoms of anxiety and depression scores, but not to types of peer engagement. Peer advocacy clients linked to HES had a longer duration of homelessness and higher rates of overall exclusion including a history of injecting drugs, incarceration, and begging, as well as more heroin use in the last year compared to non-clients.

Previous evidence from the UK is inconclusive. One trial showed peer educators did not increase uptake of tuberculosis screening, (31) while another found a positive impact on engagement with clinical hepatitis services.(15) Peer support in this trial consisted of specialist training on hepatitis C management and intensive support during and between appointments. This contrasts with the reduced service that HHPA provided due to COVID-19. The lack of reduction in DNA is disappointing, but the effect of peer advocacy on increasing inpatient admissions suggests health needs are being addressed. (32) Whether this is due to not addressing conditions earlier (there was no difference in unplanned admissions) or the timing of HHPA engagement occurring when people are symptomatic is unclear. However, the greater number of completed outpatient appointments among clients as indicated in our sensitivity analysis suggests that HHPA may be facilitating earlier treatment. Qualitative data show that HHPA adapts depending on clients’ needs and highlights the complexities of quantifying an effect.(21) Successful healthcare interactions may be linked to episodes of care and contingent on the on-going presence of the peer, while permanent changes in health care use require long-term and open-ended support. Overcoming barriers to attend inflexible appointments among this highly marginalised population, even with peer support, is complex and time-consuming.

Overall, 20% of appointments resulted in non-attendance prior to recruitment, this is almost three times higher than the national prevalence of 7.6%.(33) Most strategies and research to reduce DNA focus on changing behaviours of the individual rather than the context of healthcare. Peer advocacy challenges this by attempting to change the context of the healthcare appointment.(33) Future impact evaluations should measure longer-term changes for individuals including self-efficacy or systemic changes in healthcare. Our qualitative work also highlighted the importance of day centre and hostel settings in shaping the work of HHPA. This occurs through several mechanisms including making referrals to HHPA, ongoing staff support throughout their HHPA engagement and also as a core determinant of clients/residents’ physical and mental health. Hostel and day centre contexts vary widely depending on funding, staffing levels and burnout, issues worsened by the the pandemic’s effect on the sector. Future work also needs to explore the culture of care and support in these settings particularly in relation to stigma and the extent to which the presence of peers within a hostel, day centre of care facility may change the norms of staff and providers, reducing stigma and discrimination.

Our finding of effectiveness of peer advocacy among participants with moderate symptoms of anxiety or depression - but not severe symptoms - is in line with review evidence.(13) Further investigation on the role of peer advocacy and mental health is needed, particularly considering the high levels of anxiety and depression reported (4, 34), and to inform the growing use of psychologically-informed approaches in homeless services.(35) We urge caution that peer support is not prioritised for its lower costs rather than its specific benefits, given significant funding cuts to mental health services.

### Strengths and Limitations

The use of an outcome measure derived from hospital data is a key strength of this evaluation. Defining a comparison population was difficult given the broad criteria for HHPA engagement and is reflected in the differences observed between clients and non-clients. We tried to adjust for imbalances but there may be unmeasured confounders. We could not assume that the data were missing at random, nor did we have any validated records indicating true non-engagement with hospitals among those assigned zero records for their unlinked datasets. This could create bias in our sensitivity analysis. Sample sizes were smaller than anticipated due to fewer scheduled outpatient appointments in the follow-up period as a result of COVID-19, as well as fewer non-clients linking to HES. Clients were originally defined to be those scheduled for their first engagement with HHPA, the definition was expanded to include those who had used the service in the past 6 months due to HHPA engaging fewer new clients as a result of COVID-19.

### Conclusion

While we did not find any effect of peer advocacy on the probability of DNA at outpatient appointments, nor on the number of A&E visits, clients had more inpatient admissions. Sensitivity analysis suggested clients have more completed outpatient appointments, hospital admissions and A&E visits. Results demonstrate the importance of peer advocacy to address healthcare needs of people who are homeless and that effectiveness is linked to severity of anxiety or depression experienced by clients, information that can be used to inform scale-up of the intervention. Any certainty with which we can interpret these findings is challenged by the disruption caused to both the peer advocacy work and health services by COVID-19.

## Declarations

### Ethics

Informed consent for participation in the cohort study, the collection of personal identifiers and linking to hospital episode statistics was obtained from all participants The evaluation was approved by the London School of Hygiene and Tropical Medicine (Ref: 18021) and the Dulwich Research Ethics Committee (IRAS 271312). Data collection was performed in accordance with ESRC guidelines for working with vulnerable populations. https://www.ukri.org/councils/esrc/guidance-for-applicants/research-ethics-guidance/research-with-potentially-vulnerable-people/

### Data availability

The datasets generated and analysed during the current study are not publicly available due the inclusion of sensitive data and the potential for deductive disclosure of individuals, but are available from the corresponding author on reasonable request.

### Author Contributions

LP, AG and KB led the conception and design of the original study in collaboration with all authors. All authors acquired, analysed or interpreted the data under direction from LP and SDR. SDR curated the data and SDR and LP performed the statistical analysis with advice from EW. LP and SDR drafted the manuscript. All authors critically revised the manuscript for important intellectual content. LP is the first author and corresponding author and attest that all listed authors meeting authorship criteria and that no others meeting the criteria have been omitted.

## Acknowledgements

We would like to thank Mhonh Bancyr de Angeli, Jacqueline Craine, John Driscoll, Sam Evans, Ozgur Gencalp, Adrian Godfrey, Khalid Hussain, Mark Leonard, Stephan Morrison, Benjamin Opene, Dena Purcell, Maya Palej, Alistair Story, Keeley Tarrant, Tasia Thompson for their contributions to the study as well as all the research participants for sharing their experiences and time with us.

## References

1. Department for Levelling Up Housing & Communities. Ending Rough Sleeping For Good. London: Department for Levelling Up Housing & Communities; 2022.

2. Department for Levelling Up Housing & Communities. Official Statistics. Rough sleeping snapshot in England: autumn 2023. London: Department for Levelling Up, Housing & Communities; 2024.

3. Homeless Link Research Team. Support for single homeless people in England: Annual Review 2022. Homeless Link 2023.

4. Homeless Link. The Unhealthy State of Homelessness 2022. Findings from the Homeless Health Needs Audit. Homeless Link; 2022.

5. Lewer D, Aldridge RW, Menezes D, Sawyer C, Zaninotto P, Dedicoat M, et al. Health-related quality of life and prevalence of six chronic diseases in homeless and housed people: a cross-sectional study in London and Birmingham, England. BMJ Open. 2019;9(4):e025192.

6. Aldridge RW, Story A, Hwang SW, Nordentoft M, Luchenski SA, Hartwell G, et al. The health impact of social exclusion: a systematic review and meta-analysis of morbidity and mortality data from homeless, prison, sex work and substance use disorder populations in high-income countries. . Lancet. 2017;In press.

7. McCormick B, White J. Hospital care and costs for homeless people. Clinical Medicine. 2016;16(6):506–10.

8. Hewett N, Hiley A, Gray J. Morbidity trends in the population of a specialised homeless primary care service. British Journal of General Practice. 2011;61(584):200–2.

9. Martins DC. Experiences of homeless people in the health care delivery system: a descriptive phenomenological study. Public health nursing (Boston, Mass). 2008;25(5):420–30.

10. McNeill S, O’Donovan D, Hart N. Access to healthcare for people experiencing homelessness in the UK and Ireland: a scoping review. BMC Health Serv Res. 2022;22(1):910.

11. Omerov P, Craftman ÅG, Mattsson E, Klarare A. Homeless persons’ experiences of health- and social care: A systematic integrative review. Health Soc Care Community. 2020;28(1):1–11.

12. Schel SHH, van den Dries L, Wolf J. What Makes Intentional Unidirectional Peer Support for Homeless People Work? An Exploratory Analysis Based on Clients’ and Peer Workers’ Perceptions. Qual Health Res. 2022;32(6):929–41.

13. Lloyd-Evans B, Mayo-Wilson E, Harrison B, Istead H, Brown E, Pilling S, et al. A systematic review and meta-analysis of randomised controlled trials of peer support for people with severe mental illness. BMC Psychiatry. 2014;14(1):39.

14. Chang J, Shelly S, Busz M, Stoicescu C, Iryawan AR, Madybaeva D, et al. Peer driven or driven peers? A rapid review of peer involvement of people who use drugs in HIV and harm reduction services in low- and middle-income countries. Harm Reduction Journal. 2021;18(1):15.

15. Stagg HR, Surey J, Francis M, MacLellan J, Foster GR, Charlett A, et al. Improving engagement with healthcare in hepatitis C: a randomised controlled trial of a peer support intervention. BMC Medicine. 2019;17(1):71.

16. Surey J, Francis M, Gibbons J, Leonard M, Abubakar I, Story A, et al. Practising critical resilience as an advanced peer support worker in London: A qualitative evaluation of a peer-led hepatitis C intervention amongst people experiencing homelessness who inject drugs. Int J Drug Policy. 2021;91:103089.

17. Hewett N, Buchman P, Musariri J, Sargeant C, Johnson P, Abeysekera K, et al. Randomised controlled trial of GP-led in-hospital management of homeless people (‘Pathway’). Clinical Medicine. 2016;16(3):223–9.

18. Luchenski S, Maguire N, Aldridge RW, Hayward A, Story A, Perri P, et al. What works in inclusion health: overview of effective interventions for marginalised and excluded populations. The Lancet. 2017.

19. Barker SL, Maguire N. Experts by Experience: Peer Support and its Use with the Homeless. Community mental health journal. 2017;53(5):598–612.

20. Rathod SD, Guise A, Annand PJ, Hosseini P, Williamson E, Miners A, et al. Peer advocacy and access to healthcare for people who are homeless in London, UK: a mixed method impact, economic and process evaluation protocol. BMJ Open. 2021;11(6):e050717.

21. Guise A, Annand PJ, Hosseini P, Hudson S, Rathod SD, Platt L. Building and boosting capitals for health care access: a qualitative study of Homeless Health Peer Advocacy In London, UK. Sociology of Health and Illness (submitted). 2024.

22. Comarty H. Coronavirus: Support for rough sleepers (England). House of Commons Library HM Government; 2021.

23. Elmes J, Stuart R, Grenfell P, Walker J, Hill K, Hernandez P, et al. Effect of police enforcement and extreme social inequalities on violence and mental health among women who sell sex: findings from a cohort study in London, UK. Sexually transmitted infections. 2022;98(5):323.

24. Elwell-Sutton T, Fok J, Albanese F, Mathie H, Holland R. Factors associated with access to care and healthcare utilization in the homeless population of England. Journal of Public Health. 2017;39(1):26–33.

25. Fitzpatrick S, Johnsen S, White M. Multiple Exclusion Homelessness in the UK: Key Patterns and Intersections. Social Policy and Society. 2011;10(4):501–12.

26. Annand PJ, L. M, Marshall A, Guise A, Burrows M, Bowgett K, et al. Peer Progression: Impact of Homelessness Health Peer Advocacy on Peer Advocates. Pathways from Homelessness; 11th March Online 2021.

27. Guise A, Annand PJ, Hosseini P, Rathod SD, Platt L. Building cultural health capital to manage stigma and health care access: an evaluation of a homeless health peer support service in London, UK. British Sociological Association Medical Sociology Conference; 14th September,2022; Lancaster 2022.

28. Guise A, Burridge S, Annand PJ, Burrows M, Platt L, Rathod SD, et al. Why were COVID-19 infections lower than expected amongst people who are homeless in London, UK in 2020? Exploring community perspectives and the multiple pathways of health inequalities in pandemics. SSM Qual Res Health. 2022;2:100038.

29. Kroenke K, Spitzer RL, Williams JBW, Löwe B. An Ultra-Brief Screening Scale for Anxiety and Depression: The PHQ–4. Psychosomatics. 2009;50(6):613–21.

30. Löwe B, Wahl I, Rose M, Spitzer C, Glaesmer H, Wingenfeld K, et al. A 4-item measure of depression and anxiety: validation and standardization of the Patient Health Questionnaire-4 (PHQ-4) in the general population. J Affect Disord. 2010;122(1-2):86–95.

31. Aldridge RW, Hayward AC, Hemming S, Possas L, Ferenando G, Garber E, et al. Effectiveness of peer educators on the uptake of mobile X-ray tuberculosis screening at homeless hostels: a cluster randomised controlled trial. BMJ Open. 2015;5(9).

32. Field H, Hudson B, Hewett N, Khan Z. Secondary care usage and characteristics of hospital inpatients referred to a UK homeless health team: a retrospective service evaluation. BMC Health Services Research. 2019;19(1):857.

33. NHS England. Reducing did no attends (DNAs) in outpatient services. NHS England; 2023.

34. Rathod SD, Annand PJ, Hosseini P, Guise A, Platt L. Epidemiological features of depression and anxiety among homeless adults with healthcare access problems in London, UK: descriptive cross-sectional analysis. BJPsych Open. 2024;10(3):e93.

35. Keats H, Maguire N, Johnson R, Cockersall P. Psychologically Informed Services for Homeless People:. London: Department for Communities and Local Government; 2012.

